# Divergent Evolution in Bilateral Prostate Cancer: a Case Study

**DOI:** 10.1101/2024.08.22.24312320

**Authors:** Roni Haas, Yash Patel, Lydia Y. Liu, Rong Rong Huang, Adam Weiner, Takafumi N. Yamaguchi, Raag Agrawal, Paul C. Boutros, Robert E. Reiter

## Abstract

Multifocal prostate cancer is a prevalent phenomenon, with most cases remaining uncharacterized from a genomic perspective. A patient presented with bilateral prostate cancer. On systematic biopsy, two indistinguishable clinicopathologic lesions were detected. Whole-genome sequencing displayed somatically unrelated tumours with distinct driver CNA regions, suggesting independent origins of the two tumors. We demonstrated that similar clinicopathologic multifocal tumours, which might be interpreted as clonal disease, can in fact represent independent cancers. Genetic prognostics can prevent mischaracterization of multifocal disease to enable optimal patient management.

## Results and Discussion

In this report, we investigated the bilateral prostate cancer of a patient in his 60s, who is genetically similar to the European reference population (**Supplementary Table 1**). The patient was diagnosed with bilateral Gleason Grade (GG) 1 tumours on systematic biopsy and was upgraded to GG3 in both on pathology from surgical specimens. Both tumours harboured 60% pattern 4 and 4% tertiary pattern 5. There were no cribriform or intraductal components and both tumors were confined to the prostate. The uncanny clinical and histologic similarities between the two tumors (**Figure 1A**) inspired molecular evaluations to test if nearly indistinguishable clinicopathologic features reflect similar tumor biology.

**Figure 1.**
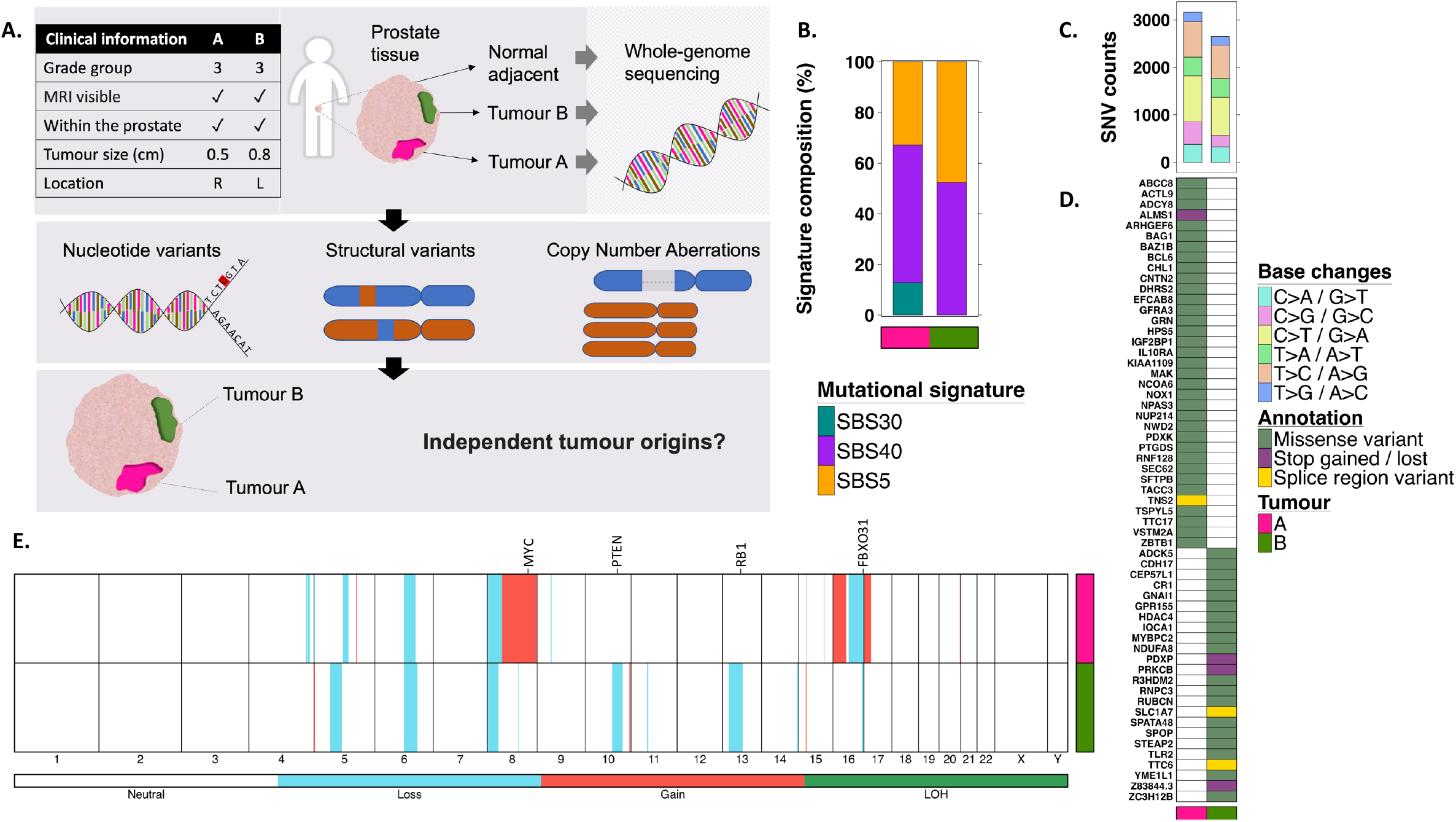
Genomic landscape. **A**. Schematic of study design and workflow. **B**. Mutational signatures. Signature etiologies: SBS30, defective DNA base excision repair due to NTHL1 mutation; SBS40, unknown; SBS5, unknown. **C**. SNV counts categorized by the base change type. **D**. Functionally affected genes by SNVs **E**. Genome-wide view of CNAs. Chromosome numbers are presented on the x-axis. The colored lines represent CNA classes. LOH, loss of heterozygosity.

We sampled the separate tumours (A and B) from freshly frozen radical prostatectomy tissue. A benign prostate tissue, procured simultaneously from the same patient, was used as a reference control. DNA whole genome sequencing (WGS) was carried out to interrogate the genomic landscape and evolutionary relationship of the tumors (**Figure 1A**; **Supplementary Figure 1**).

We first evaluated the extent of genomic instability in each tumour as reflected by mutation counts. Larger numbers of somatic single nucleotide variants (SNVs) and structural variants (SV) were detected in tumour A compared to B, and none were shared between the two (**Table 1; Supplementary Tables 2-3; Supplementary Figure 2**). Despite this, SNVs in both tumours were largely the consequences of the same mutagenic processes (**Figure 1B**). The tumors experienced similar profiles of base change mutations (**Figure 1C)** and contained multiple SNVs that are predicted to have a significant protein functional effect. No genes shared functional SNV in both tumours (**Figure 1D)**. Among driver genes, *BCL6* and *NUP214* were functionally affected in Tumour A, and *SPOP* in Tumour B.

**Table 1.**
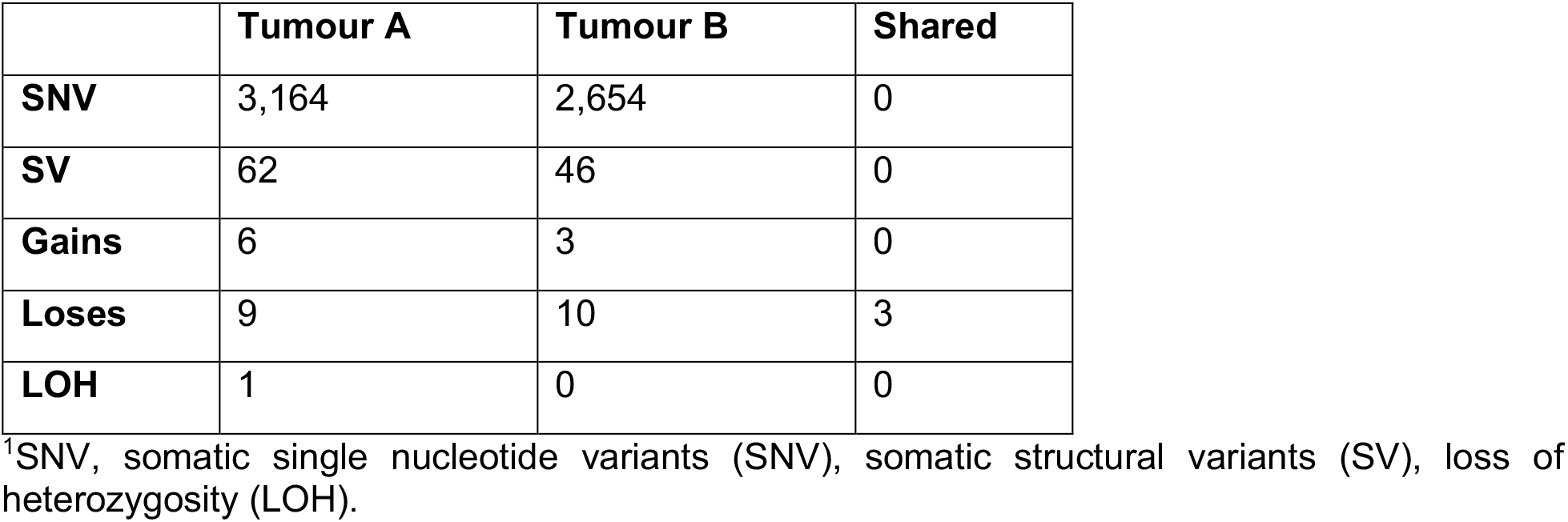
mutation counts.

Next, we compared the copy number aberration (CNA) profiles of tumours A and B to identify possible shared CNA-driver regions, which could imply a shared evolutionary origin (**Table 1; Figure 1E; Supplementary Table 4**). Three CNA regions overlapped (**Supplementary Figure 3**), but only one, on chromosome 16, was a CNA-driver region spanning *FBXO3* (**Supplementary Figure 3C**). However, the breakpoints of these events differ, indicating convergent evolution rather than a shared origin.

Consistent with SNV and SV results, slightly higher genomic instability originating from CNAs was observed in Tumour A (**Table 1**). This involved CNA-driver regions, including *MYC* and *NCOA2* gains on chromosome 8q and *NKX3-1* deletion on chromosome 8p, typically seen in prostate cancer. By contrast Tumour B was characterized by *PTEN* deletion on chromosome 10, and *RB1* deletion on chromosome 13. Overall, the CNA landscape displayed distinctive clonal drivers.

Taken together, our results support distinct genetic origin of the two tumours, indicating that they arose independently and represent two separate cancers. The remarkably similar clinicopathologic features could have been mistakenly considered clonal without further genetic characterization. This study, and previous works^1–3^ emphasize that the genetic prognostic should be carefully considered in situations of multifocal prostate tumors, which are relatively common^4,5^. Mischaracterizing multifocal lesions as a single prostate cancer may prevent optimal patient management.

## Methods

### Sample preparation and WGS

Two tumor foci, A and B were punched out from fresh OCT embedded tissue blocksafter central pathology assessment. Tumor DNA was extracted from the same tumor tissue cores using Qiagen All Prep DNA mini kit (Qiagen, cat. 80204). The sample from Tumor A was in the anterior right, and from Tumor B in the posterior left, close to the dominant tumor (0.8 cm). A normal prostate core was sampled from a distal OCT block, from which the normal DNA was extracted using Qiagen QIAamp DNA mini kit (Qiagen, Cat 51304).

AnaPrep Tissue DNA Extraction kit (BioChain, cat. Z1322004) was used for DNA isolation. 500 to 1000 ng of input DNA was used for the library construction. Libraries were prepared using KAPA HyperPrep kit (Cat. KR0961, Roche). High throughput sequencing was performed by Illumina NovaSeq 6000 sequencer S4 platform with 150bp paired-end reads in 1.5 lanes. Coverage depths of 120x for the tumor samples and 80x for the normal tissue were achieved. A data quality check was done on Illumina SAV. Demultiplexing was performed with Illumina Bcl2fastq v2.19.1.403 software.

### WGS processing

DNA reads were tested with FastQC v0.11.8^6^ for quality assurance before alignment. The reads were then mapped to the human GRCh38 reference genome using BWA-MEM2 v2.2.1^7^, and aligned SAM files were converted to BAM files using SAMtools v1.12^8^. Next, using Picard Tool’s v2.26.10^9^, the resulting BAM files were sorted in coordinate order, duplicates were marked, and the BAM files were indexed. The indexed BAM files went through indel-realignment using the Genome Analysis Toolkit (GATK) v3.7.0^10^ and base quality score recalibration was done using GATK v4.1.9.0^10^ in tumour-normal pairs. Separate BAM files were generated for the tumour and normal samples, and their headers were modified using SAMtools v1.12^8^. Qualimap version v.2.2.2-dev^11^ was used to verify high coverage across the genome (**Supplementary Figure 1**). The mean coverage was 105X for T1, and 95X for T2.

### Mutation calling

Processed BAM files from the previous step were taken for mutation calling in tumour-normal pairs. SNVs were called separately using three algorithms, Mutect2 of GATK v4.2.0.0^10^, Strelka2 v2.9.10^12^ and SomaticSniper v1.0.5.0^13^. To increase reliability, SNVs were declared only for sites that were detected using at least two algorithms for each tumour site. Strict germline variant filtering was performed, excluding any variant in the normal control sample and in gnomAD v3.1.2^14^. The called SNVs were annotated using SnpEff v5.0e^15^. Mutational signatures were identified using SigProfilerExtractor v1.14^16^.

SVs were identified using DELLY v0.8.7^17^. For reliability, detected sites were filtered to exclude germline structural variants. SVs that were identified with DELLY, despite very low coverage in their area in the normal sample, were excluded. Annotations were obtained using SnpEff v5.0e^15^. CNAs were detected using Battenberg v2.2.9 ^18^, with a purity of 0.41 and 0.21 for tumours A and B respectively, and ploidy of ∼2 for both. Each call was reviewed manually to filter out false findings.

### Genetic ancestry

Genetic ancestry was predicted using Peddy v0.4.8, with WGS of the normal sample as input^19^. The prediction results calculated by Peddy are presented in **Supplementary Table 1**.

### Visualization

Visualizations were generated in the R environment v4.2.0 using the BoutrosLab.plotting.general package v7.0.3^20^.

## Author Contributions

Sample preparation: R.R.H

Data processing analyses and visualization: R.H

Generated tools: Y.P, L.Y.L., T.N.Y, R.A

Clinical annotation: A.W., R.E.R. Supervised research: R.E.R, P.C.B

Wrote the first draft of the manuscript: R.H

Approved the manuscript: all authors.

## Ethical Statement

The Institutional Review Board of the University of California Los Angeles gave ethical approval for this work. The patient signed a universal consent for analyses of biological samples (#15-001395).

## Data and Materials Availability

All data produced in the present study are available upon reasonable request to the authors.

## Acknowledgments

R.H is supported by EMBO Postdoctoral Fellowship ALTF 1131-2021 and the Prostate Cancer Foundation Young Investigator Award 22YOUN32. R.A is supported by the NIH grant T32GM008042. ABW was supported by the UCLA Dr. Allen and Charlotte Ginsburg Fellowship in Precision Genomic Medicine and the Prostate Cancer Foundation Young Investigator Award (23YOUN21). This work was supported by the NIH through awards P30CA016042, U2CCA271894, R01CA270108 and P50CA092131. It was supported by the DOD through awards W81XWH2210247 and W81XWH2210751. This work was supported by a Prostate Cancer Foundation Special Challenge Award to PCB (Award ID #: 20CHAS01) made possible by the generosity of Mr. Larry Ruvo.

