## Supplementary Figures for "Divergent Evolution in Bilateral Prostate Cancer: a Case Study"

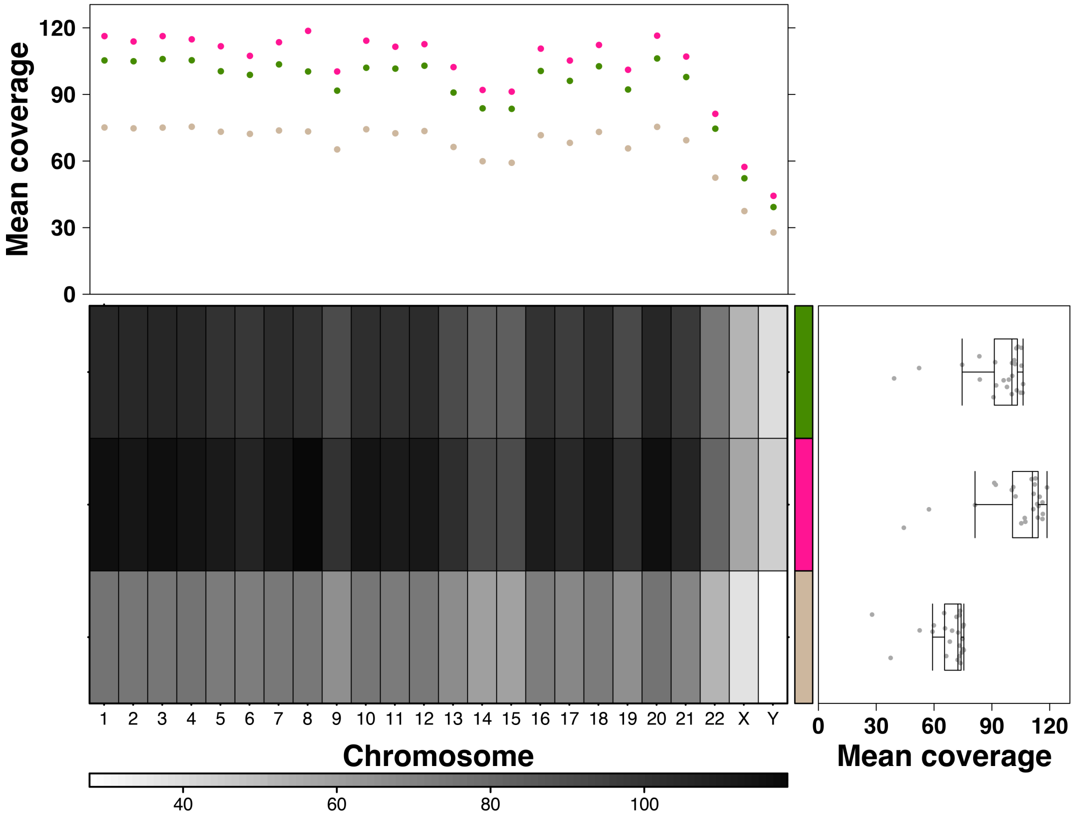


**Supplementary Figure 1. Read coverage across the genome.** Chromosome numbers are presented on the x-axis. The heatmap colors show the read coverage. Pink, tumour A; Green, tumour B, Tan, normal sample.

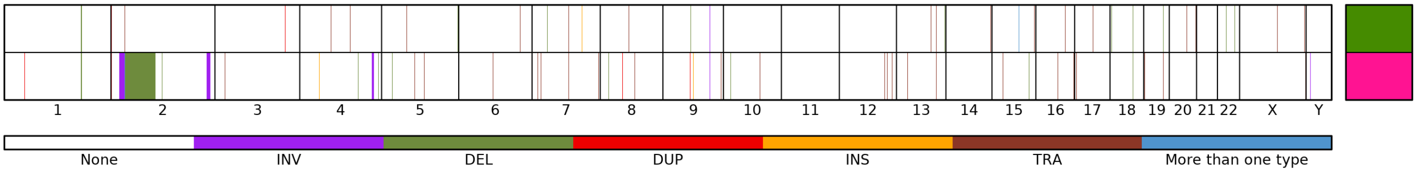


**Supplementary Figure 2. The SV profiles of Tumours A and B.** Chromosome numbers are presented on the x-axis. The colored lines represent types of SVs: INV, inversion; DEL, deletion; DUP, duplication; TRA, translocation. Pink, Tumour A, Green Tumour B.

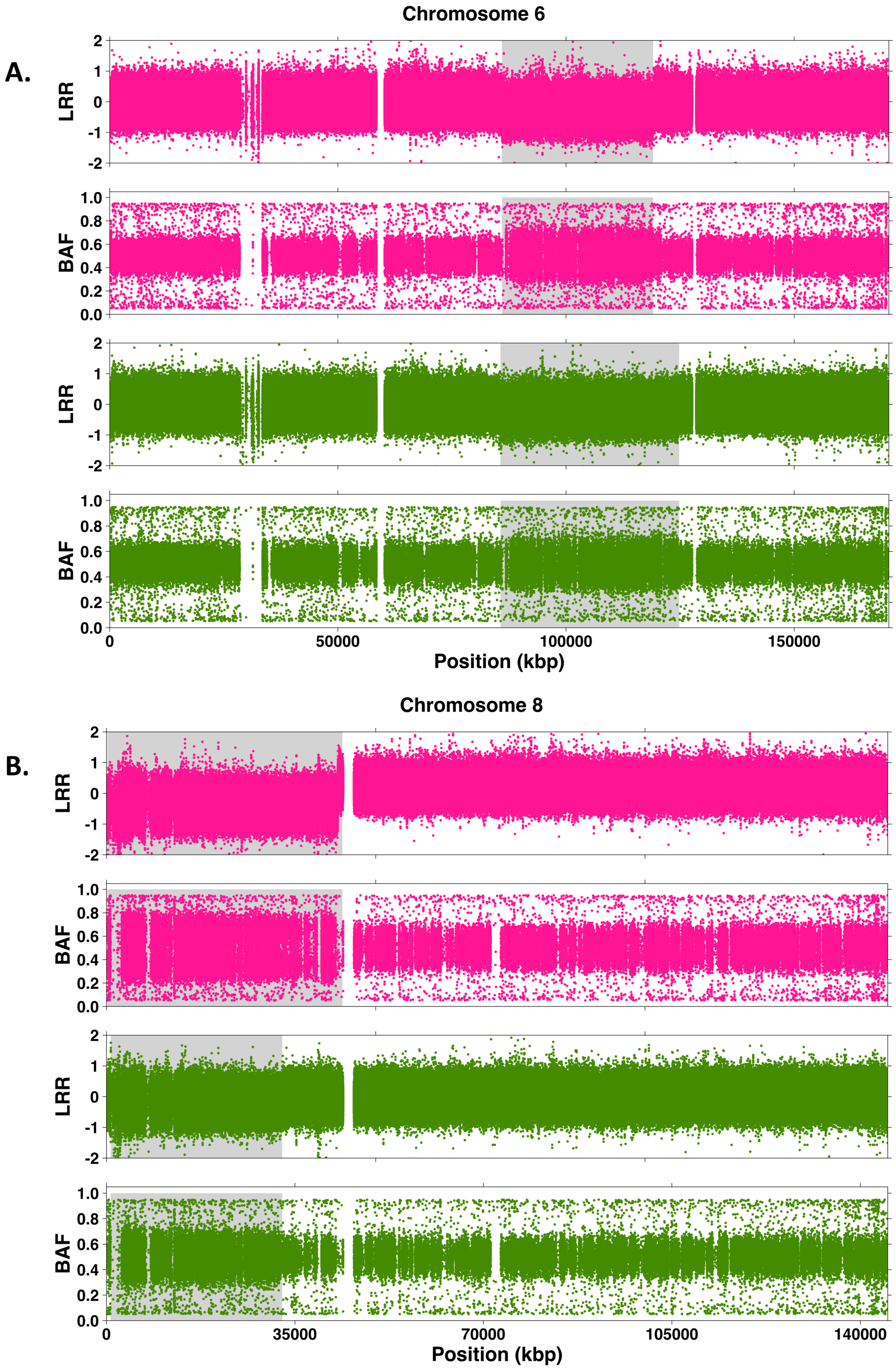


**Supplementary Figure 3. Shared CNA regions.** Presented are CNA regions that show similar log R ratio (LRR) and B- Allele frequencies (BAF) patterns between Tumour A and B in chromosomes 6 (**A**), 8 (**B**) and 16 (**C**). Homozygous SNP (BAF > 0.95 or BAF < 0.05) were excluded for visualization. Pink, Tumour A; Green, Tumour B. Marked in grey are the CNA regions.

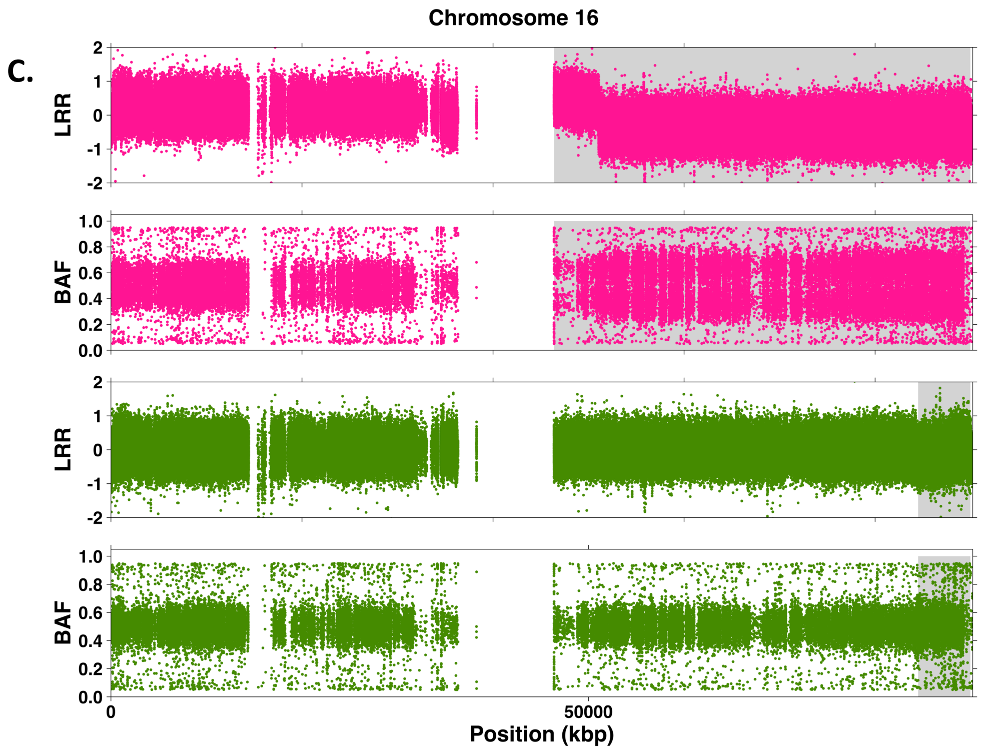


**Supplementary Figure 3. Shared CNA regions.** Presented are CNA regions that show similar log R ratio (LRR) and B- Allele frequencies (BAF) patterns between Tumour A and B in chromosomes 6 (**A**), 8 (**B**) and 16 (**C**). Homozygous SNP (BAF > 0.95 or BAF < 0.05) were excluded for visualization. Pink, Tumour A; Green, Tumour B. Marked in grey are the CNA regions.
